# The Silver Trauma Review Clinic: A novel model of care to manage non-operative injuries in older patients

**DOI:** 10.1101/2023.02.27.23286493

**Authors:** H Smyth, D Breslin, L Mullany, V Ramiah, R Riches, R Laguna, P Morgan, C Byrne

## Abstract

**Background:** Increasing numbers of older patients are presenting to emergency departments(ED) following trauma. These patients require multidisciplinary care that the traditional trauma model fails to provide. A Silver Trauma Review Clinic(STRC) was developed in conjunction with the geriatric, ED and multidisciplinary services to improve the post-discharge care of patients with non-operative traumatic injuries.

We aimed to assess the STRC by reviewing the journey and outcomes of patients who attended the clinic and examining new diagnoses and interventions.

**Methods:** A retrospective review of electronic chart data was performed on all patients who attended the clinic over the initial 1 year period.

**Results:** 137 patient were reviewed with a median age of 80(IQR 12.5), 69% female. The median clinical frailty scale was 3 with a median time from the patient’s initial ED presentation to clinic of 15 days(IQR 11.25) and median time from initial review to discharge 20 days(IQR 34). 71% of presentations were as a result of falls under 2 metres. Primary injuries were 34% vertebral fractures, 45% limb fracture, 18% thoracic trauma, 11% pelvic trauma with 15% of patients suffering from multiple injuries. Patients attending the STRC had a comprehensive geriatric assessment with abnormal Mini-Cog assessments found in 29%, a new diagnosis of osteoporosis in 43% and orthostatic hypotension diagnosed in 13% of patients. 61% were discharged to primary care, 19% linked into a specialist geriatric clinic.

**Conclusion:** The STRC is a novel approach allowing timely, patient focused, comprehensive and collaborative trauma care of older patients following non-operative injuries.

## Introduction

Management of trauma in older adults, or “silver trauma”, can be challenging due to pre-existing co-morbidities and frailty [1,2]. Relatively minor injuries can have a significant impact on functional outcome [3,4,5]. While many injuries will not require hospital admission, patients can experience ongoing issues with medication management, side effects, undiagnosed or missed injuries and pain [2,4,5]. Previous studies have established the positive effects of geriatrician review on long-term outcome following trauma [6,7,8,9]. The average age trauma patients in Ireland is increasing, with the most recent data from the National Office of Clinical Audit recording a mean age of 61 for major trauma patients [10].

We sought to improve post-discharge care for patients who attended the Emergency Department (ED) with non-operative injuries. These were patients who did not require admission and who had sustained injuries that were amenable to conservative management – either due to the injury pattern itself, or patient factors, such as co-morbidity, frailty, or baseline function. Prior to our intervention, these injuries were managed in the orthopaedic fracture clinic which lacks access to specialist geriatric care and does not provide services such as cognitive screening and falls risk assessment. While patients could also be referred to a specialist falls clinic or geriatric clinic, this was at the discretion of the referring emergency practitioner and occurred on an ad-hoc basis. We therefore recognised a deficit in the post-discharge care for these patients, and sought to address this issue.

To this end, a Silver Trauma Review Clinic (STRC) was designed and commenced in May 2021. The purpose of the clinic is to provide comprehensive multidisciplinary management for older patients in a timely manner following trauma. We aimed to assess this by reviewing the journey and outcomes of patients who attended the STRC and examining new diagnoses and interventions arising from the clinic.

## Methods

### Setting

The STRC is based in a level 4 teaching hospital in Dublin’s inner city. In 2021 the hospital was selected as the designate Major Trauma Centre for the Central Trauma Network in Ireland. The STRC clinical team consists of a consultant in emergency medicine (with a special interest in Geriatric Emergency Medicine), a consultant geriatrician, a physiotherapist and an advanced nurse practitioner (ANP).

### Clinic Eligibility

The clinic reviews patients >= 65 years of age with a Clinical Frailty Scale (CFS) >2, who are discharged from the ED following a trauma, or with a non-operative fracture. These include non-operative fragility fracture, e.g. fractures of distal radius, pelvis or vertebral column, soft tissue injuries and minor head injuries/concussion.

### Processes and Design

Patients are referred to the STRC by an ED clinician or by the hospital’s Frailty Intervention Team at the time of ED review. Other suitable patients are recruited by reviewing referrals made to the orthopaedic fracture clinic on a secure medical messaging app. The referrals to fracture clinic are monitored by consultant orthopaedic surgeons and by members of the STRC, allowing identification and diversion of suitable patients to the STRC. The clinic is held for a half-day each week. On average, patients are seen in the clinic approximately 2 weeks after their initial trauma.

The STRC aims to evaluate patients and assess for occult injury, identify medical complications following injury, perform falls risk and bone health assessments and to develop a plan for rehabilitation. A standardised electronic document is used for each patient (Appendix A). Patients attending for the first time are assessed by each member of the clinical team and a multidisciplinary management plan is created. The consultant in emergency medicine is responsible for conducting a tertiary survey and for management of the patient’s injuries, but pathways have been developed to access orthopaedic input via fracture clinic and vertebroplasty via interventional radiology, if indicated.

Patients are often seen again 2-4 weeks later to ensure bone healing, review pain and function and discuss investigation results. On discharge from the clinic, the completed assessment document is sent to the patient’s general practitioner (GP). Patients may be discharged to their GP, physiotherapy, geriatric review clinic or referred for further specialist review.

### Data Collection / Analysis

A retrospective electronic chart review was performed on all patients who attended the clinic from 1^st^ June 2021 to 1^st^ June 2022. Data was collected from the electronic assessment document completed by each member of the MDT during each patient review. The data collection was performed by clinicians who were not involved in establishing the clinic, or directly involved in patient care in the clinic. Anonymised data was collected and simple summary statistics were used to describe the patients attending the clinic.

### Patient and Public Involvement

Patients were not involved in the design or implementation of this study.

## Results

Over a 1 year period, 161 patients were referred to the STRC. 9 (6%) patients did not attend and 15 (8%) patients are awaiting further investigations or reviews (e.g. scans, bone health review).

Table 1 describes the baseline characteristic and injuries of patients attending the STRC. In total 137 patients were fully reviewed with a median age of 80 (IQR 12.5) and 69% female (n=95). The median CFS was 3 with 75 patients (55%) having a CFS of 1, 2 or 3 indicating patients are respectively very fit, fit or managing well. 27 (20%) patient had a diagnosis of dementia with other co-morbidities listed in Appendix B.

**Table 1.** Characteristics, injuries & timelines of patients attending the STRC

|  | Median (IQR) or n (%) |
| --- | --- |
| **Total number** | 137 |
| **Age (years)** | 80 (12.5) |
| **Female** | 95 (69%) |
| **Dementia** |  |
| **Osteoporosis** | 27 (20%) |
|  | 42 (31%) |
| **Medications** |  |
| **0-5 tablets** | 56 (41%) |
| **6-10 tablets** | 54 (39%) |
| **>11 tablets** | 24 (18%) |
| **Clinical frailty scale** |  |
| **1** | 14 (10%) |
| **2** | 27 (20%) |
| **3** | 34 (25%) |
| **4** | 26 (19%) |
| **5** | 22 (16%) |
| **6** | 8 (6%) |
| **7** | 4 (3%) |
| **Unknown** | 2 (1%) |
| **Functional aid** |  |
| **None** | 77 (54%) |
| **Walking stick** | 29 (21%) |
| **Crutches** | 4 (3%) |
| **Zimmer frame** | 13 (9%) |
| **Rollator** | 8 (6%) |
| **3 wheeled rollator** | 9 (7%) |
| **Wheelchair** | 0 (0%) |
| **Timelines** |  |
| **Injury to presentation (days)** | 2 (6) |
| **Presentation to clinic (days)** | 15 (11.25) |
| **Review to discharge (days)** | 20 (34) |
| **Mechanism of injury** |  |
| **Fall < 2m** | 98 (72%) |
| **Fall > 2m** | 12 (9%) |
| **Road traffic accident** | 4 (3%) |
| **Assault** | 1 (0.7%) |
| **Atraumatic** | 21 (15%) |
| **Primary injuries** |  |
| **Limb fracture** | 62 (45%) |
| **Vertebral fracture** | 48 (35%) |
| **Thoracic injury** | 24 (18%) |
| **Pelvic fracture** | 15 (11%) |
| **Head injury** | 10 (7%) |
| **Soft tissue injury** | 4 (3%) |
| **Tertiary survey injuries** |  |
| **Limb fracture** | 11 (8%) |
| **Vertebral fracture** | 8 (6%) |
| **Thoracic injury** | 5 (4%) |
| **Head injury** | 1 (0.7%) |
| **Pelvic injury** | 0 (0%) |

Median time from initial presentation to ED to review was 15 days (IQR 11.25) and median length of time from initial review to discharge was 20 days (IQR 34). The majority of patients required 1 or 2 reviews prior to discharge from the clinic (n=120, 88%).

71% of patients’ presentations (n=97) to the STRC were caused by a fall from less than 2m. 15% of patients (n=15) had atraumatic presentations (fragility fractures) with other mechanisms of injuries and injuries summarised in Table 2. Primary injuries were limb fractures (n=62, 45%), vertebral fractures (n=47, 34%), thoracic injuries (n=24, 18%), pelvic fractures (n=15, 11%), head injuries (n=10, 7.2%) or soft tissue injuries (n=4, 3%). Tertiary survey in the STRC identified previously unrecognised injuries in 24 patients (18%) following review. In total, 56 patients were reviewed with vertebral fractures. 87% of these patients (n=49) were further investigated with a CT or MRI and 95% of patients (n=53) were referred for physiotherapy (Appendix C).

**Table 2.**
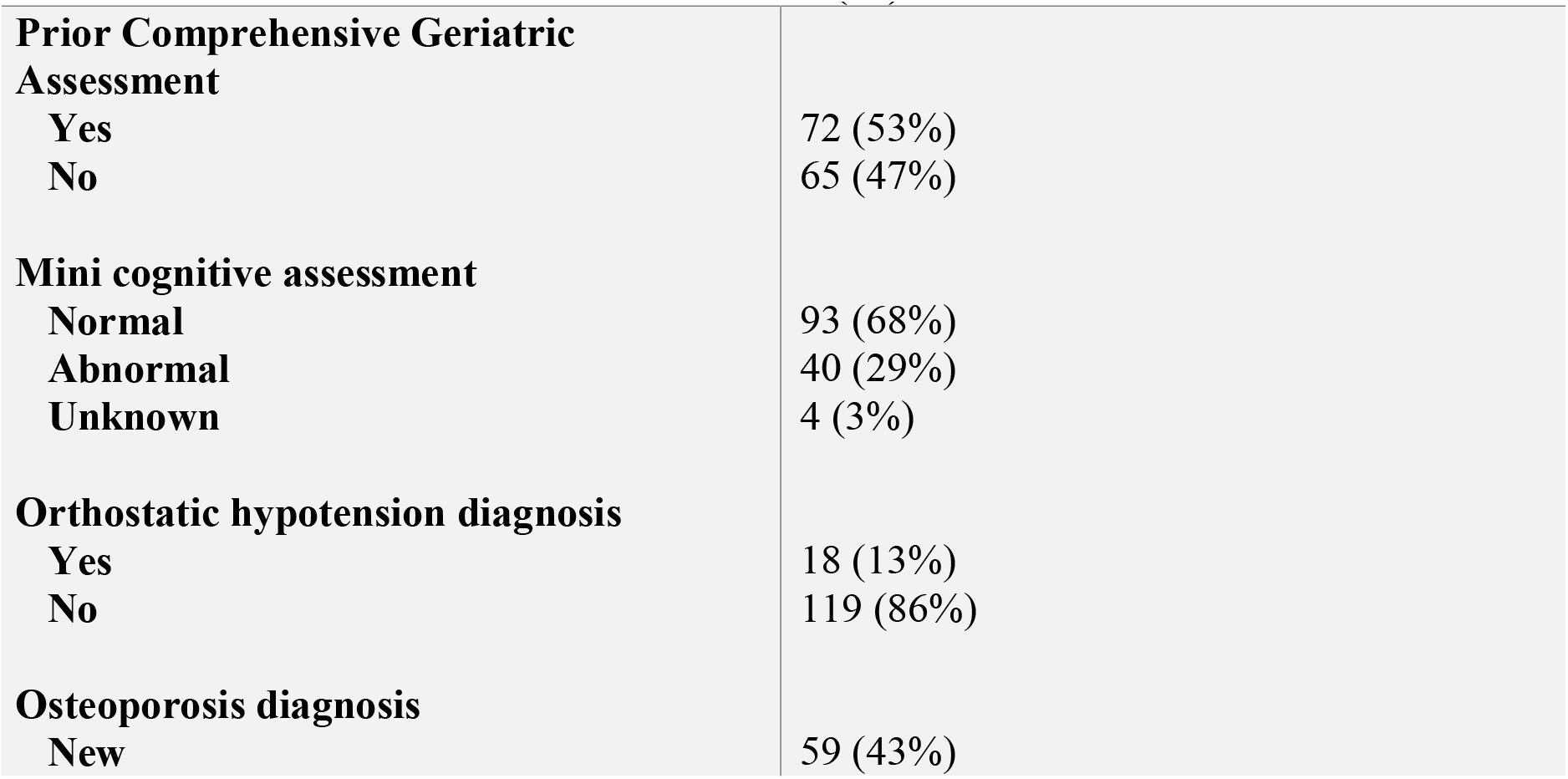
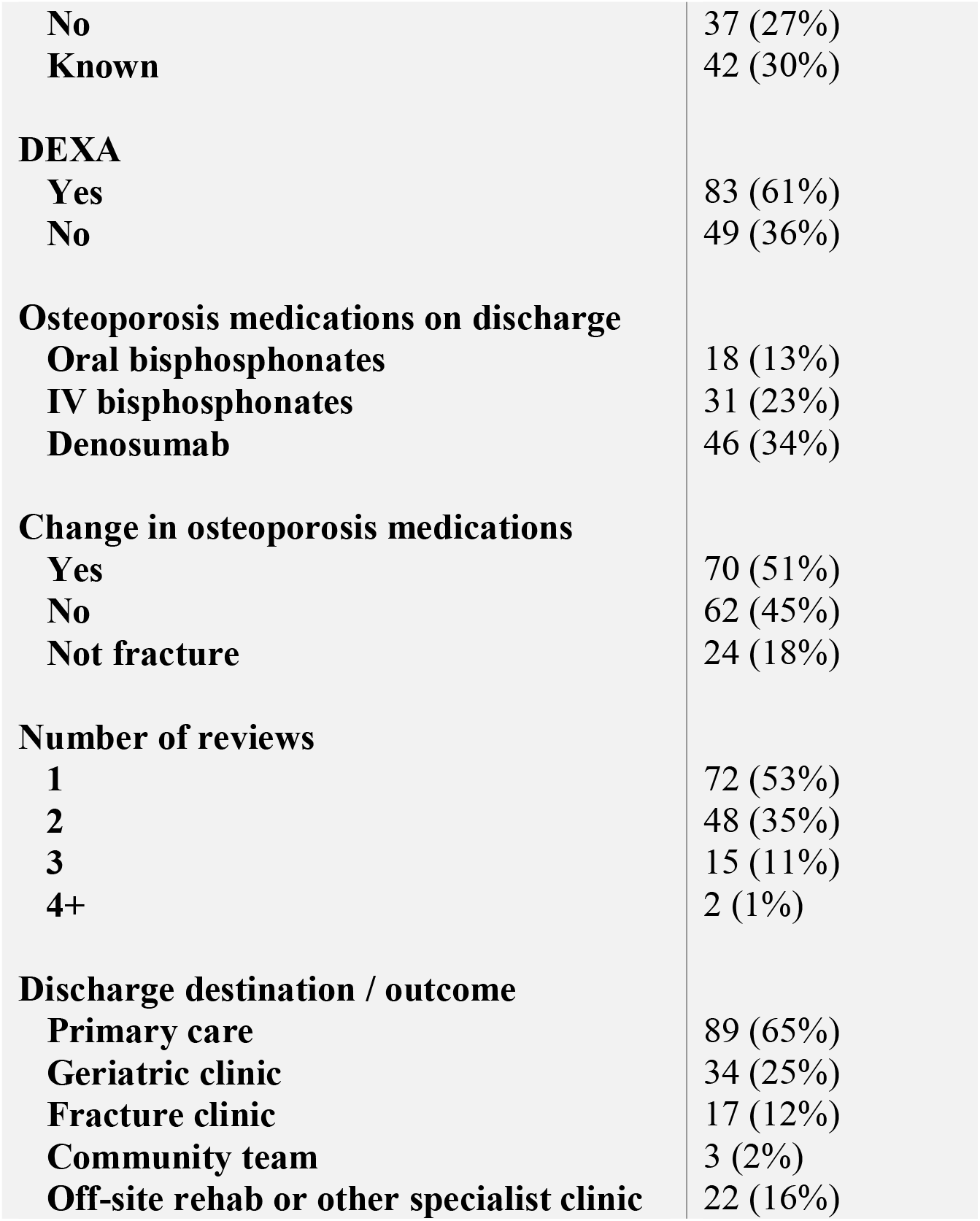
Assessments and outcomes of patients attending STRC

72/137 (53%) of patients attending the STC had a full comprehensive geriatric assessment (CGA) carried out prior to their attendance in STRC. All patients in STRC underwent a CGA as detailed by the Silver Trauma Assessment document (Appendix A).

Table 2 summarises the assessments and outcomes of patients attending the STRC. Abnormal Mini-Cog assessments were found in 40 patients (29%). Of these, 22 patients had a prior diagnosis of neurodegenerative disorder / cognitive impairment. 18 patients had a newly identified abnormal mini-cog – of these, 3 required referral inpatient rehabilitation (under the care of a geriatrician). All others were referred to the medicine for the older person clinic for further evaluation (under the ongoing care of the geriatrician who reviewed them in the STRC).

A diagnosis of orthostatic hypotension was made in 18 patients (13%).

A new diagnosis of osteoporosis was made in 59 patients (43%) with 42 patients (31%) having known osteoporosis. All 42 of the patients with a recorded diagnosis of osteoporosis were already prescribed bone protection medication. 28 patients reported compliance with prescribed bone protection medication, 8 were non-compliant, and for 6 patients compliance was not recorded. Of the 42 patients with a prior diagnosis of osteoporosis, 13 had their bone protection medication changed in the clinic.

83 patients (61%) had a DEXA scan in the STRC and half of patients had a change in their osteoporosis medications. 31 patients (23%) were linked into to an IV zoledronic acid clinic.

Overall, 84 patients (61%) were discharged to primary care, 26 patients (19%) were linked into a specialist geriatric clinic for follow up, 15 patients (11%) required further follow up in a fracture clinic. 4 patients (3%) were linked in with specialist geriatricians in the community and 22 patients (16%) were referred to off-site rehabilitation or other specialist clinics.

## Discussion

Establishing a Silver Trauma Review pathway and clinic for patients >65 presenting to the ED provides a unique opportunity for follow up, diagnosis and comprehensive review of older patients following a trauma. This dedicated outpatient service allows timely identification of important health, mobility and functional issues. Patients reviewed in the STRC were predominantly older (median age 80) and female.

Half had a CFS of 4 or more, reflecting a potential vulnerable or frail cohort of patients [11]. As previously shown, these patients highly benefit from CGA to identify, coordinate and treat their needs [12]. 30% of patients in our review had a clinical frailty score of 1 or 2 on review in the STRC, and therefore did not meet the criteria for referral to the STRC. However, these may be patients who presented to the Emergency Department with a frailty syndrome, and further study and review of the referral criteria may determine which patients receive maximal benefit from this novel service.

One of the most important aspects of care offered by the STRC is combined, multidisciplinary specialist assessment by a consultant geriatrician, an emergency medicine physician, a physiotherapist and an ANP. This facilitates a comprehensive evaluation and review of the factors contributing to emergency presentations, such as medications, mobility, bone health, cognition and falls assessments. On screening when indicated, abnormal cognitive assessments were picked up in about 1/3^rd^ of patients and appropriate education and follow up could be arranged. Orthostatic hypotension was diagnosed in a small number of patients. The STRC facilitates a more in-depth and holistic analysis of the patients’ primary concerns such as pain, mobility, fear of falling, lifestyle and possible future planning.

Due to their complexity and reduced physiological reserves, older patients may have atypical presentations [13,14]. Thanks to the short follow-up times, the STRC provides an opportunity for tertiary trauma surveys, with occult or further injuries identified in nearly 1/5 of patients reviewed. The STRC allows quick access to imaging. With access to DEXA scans and bone heath screening, new osteoporosis was diagnosed in 43% of our patients and half of our patients had changes made to their osteoporosis treatment to improve compliance and prevent further fractures.

Outpatient combined comprehensive specialist care has been previously described in hip fracture patients with the benefit of identifying and managing issues overlooked during the patients’ acute care [8]. Similarly, combined specialist and geriatric care through orthogeriatrics, oncogeriatrics, geriatric cardiology is emerging with positive outcomes for an increasing older population requiring mixed patient-centred skills [15,16, 17].

Strengths of our study include data collection from a single electronic clinic template completed by each member of the interdisciplinary team on review. Our centre has been identified as the regional Major Trauma Centre and this model of combined outpatient specialist care of older patients could be a future standard of care for the management of older patients with non-operative trauma.

Limitations include selection bias of patients. Our review included a high proportion of patients who were not frail. However, this is a unique opportunity for this cohort of non-frail patients to be given preventive lifestyle advice on “healthy ageing” including physical activity, socialising, good nutrition [18]. The generalizability of this study is also limited as it is a single site study and the potential benefits of STRC are yet to be proven through prospective validation. Future research will assess the impact of the STRC on functional outcomes, admission avoidance and surveys on patient and health care satisfaction.

## Conclusion

The STRC is a novel model of care allowing review of older patients with nonoperative trauma following presentation to ED. The short time follow up allows a focused comprehensive and collaborative multidisciplinary assessment addressing primary and secondary injuries, potential further investigations and treatment. Following a CGA, there is also a unique opportunity to diagnose geriatric and frailty syndromes while linking patients into appropriate specialty and community services.

## Data Availability

All data produced in the present study are available upon reasonable request to the authors.

## Contributorship statement

All authors take responsibility for the content of the work submitted and have helped draft & review the manuscript.

HS: Conception or design of the work. Data collection. Data analysis and interpretation. Drafting the article. Critical revision of the article. Final approval of the version to be published.

DB: Conception or design of the work. Drafting the article. Critical revision of the article. Final approval of the version to be published.

LM: Data collection. Data analysis and interpretation. Drafting the article. Final approval of the version to be published.

VR: Conception or design of the work. Critical revision of the article. Final approval of the version to be published.

RR: Conception or design of the work. Critical revision of the article. Final approval of the version to be published.

RL: Conception or design of the work. Critical revision of the article. Final approval of the version to be published.

PM: Conception or design of the work. Critical revision of the article. Final approval of the version to be published.

CB: Conception or design of the work. Data analysis and interpretation. Drafting the article. Critical revision of the article. Final approval of the version to be published.

## Data Sharing Statement

All data relevant to the study are included in the article or uploaded as supplementary information

## Ethical approval statement

Ethical exemption and approval for external dissemination was provided by the Clinical Audit and Effectivenes Committee, Mater Misericordiae University Hospital. Ref. CA22-087

