## Supplemental File A for "The Silver Trauma Review Clinic: A novel model of care to manage non-operative injuries in older patients"

**Silver Trauma Review Clinic**
**Patient Name:** **MRN :** **Date:**
**Assessor name and designation:**

| TRAUMA ASSESSMENT | |
|---|---|
| **Name:**<br>**MRN:**<br>**Address:**<br><br>**DOB:**<br>**Phone numbers:**<br>**NOK Ph:**<br>**GP:** | **Date of Injury:**<br><br>**MECHANISM OF INJURY:** |
| **INJURIES: PRIMARY ASSESSMENT** | **INJURIES SECONDARY ASSESSMENT** |
| 1.<br>2.<br>3.<br>4.<br>5.<br>6. | 1.<br>2.<br>3.<br>4.<br>5.<br>6. |
| **Past Medical history:** | **Medications:** |
| **Imaging:** | **Issues:** |

| INVESTIGATIONS REQUIRED |
|---|
| Bloods: Geriatric Screen **Y** ☐ **N** ☐<br>Imaging: Further imaging required: **Y** ☐ **N** ☐<br>Other: ECG **Y** ☐ **N** ☐ |

**Silver Trauma Review Clinic**
**Patient Name:** **MRN :** **Date:**
**Assessor name and designation:**

| | **CURRENT ISSUES** | |
|---|---|---|
| **Pain:** | Location, aggravating factors, analgesia<br>Pain score: 0 ☐ 1 ☐ 2 ☐ 3 ☐ 4 ☐ 5 ☐ 6 ☐ 7 ☐ 8 ☐ 9 ☐ 10 ☐ | |
| **Medications** | Paracetamol Y ☐ N ☐<br>Codeine Y ☐ N ☐<br>NSAIDs Y ☐ N ☐<br>Opiates Y ☐ N ☐<br>Neuropathic Y ☐ N ☐<br>Topical Agents Y ☐ N ☐<br>Capsaicin Y ☐ N ☐<br>Pain patches Y ☐ N ☐ | Sleeping tablets Y ☐ N ☐<br>Heat/Ice Y ☐ N ☐<br>TENS Y ☐ N ☐<br>Others: |
| **Psychosocial/Mood:** | Do you feel depressed or low? Y ☐ N ☐<br>Flashbacks ☐ Night terrors ☐ | Are you lonely/isolated? **Y** ☐ **N** ☐ |
| **Sleeping Pattern** | Disturbed Y ☐ N ☐<br>Slightly < 1h ☐ Mild 1-2h ☐ Moderate 2-3h ☐ Severe 3-4h ☐ Completely > 4 h ☐ | |
| **Recent behaviour change/Agitation/Aggression** | Y ☐ N ☐ | |
| **Constipation** | Y ☐ N ☐<br>Laxatives Y ☐ N ☐ | |
| **PLAN** | | |

| | |
|---|---|
| Pain Management | Imaging |
| Physiotherapy ☐<br>Plastic Surgery ☐<br>Orthopaedics ☐<br>Thoracic Surgery ☐<br>Other ☐ | **FOLLOW UP:**<br>Review in STRC ☐ When?<br>RAC ☐<br>COTOP Day Ward ☐<br>PCT ☐<br>Fracture clinic ☐ |

**Silver Trauma Review Clinic**
**Patient Name:** **MRN :** **Date:**
**Assessor name and designation:**

### BONE HEALTH

| | | |
|---|---|---|
| Weight: kg<br>Height: cm<br><br>Smoking: **Y** ☐ **Ex** ☐ **N** ☐<br>**Alcohol: units/week**<br><br>**Dairy intake:**<br><br>**Calcium/Vit D Supplementation:** | Previous fracture Y☐ N☐<br>If yes: location?<br><br>1st degree family history of hip fracture or osteoporosis? Y☐ N☐<br><br>**Age of Menarche:**<br>**Age of Menopause:**<br><br>**Current Treatment:**<br><br>**Compliance:** | **Secondary Osteoporosis**<br>Long term steroid use. Y☐<br>T1DM Y☐<br>Hyperthyroidism Y☐<br>Cushing's Disease Y☐<br>Coeliac or absorption disorder Y ☐<br>IBD Y☐<br>CKD Y☐<br>Eating disorder Y☐<br>Chronic Liver disease Y☐<br>Rheumatoid Arthritis Y☐ |

### DEXA

Previous DEXA Scan results :

### FRAX Score

https://www.sheffield.ac.uk/FRAX/tool.aspx?country=48

| Cognitive Screen | Clinical Frailty Scale |
|---|---|
| **Mini Cog** Normal ☐ Abnormal☐ | **CFS Score:** |

### GERIATRICIAN ASSESSMENT

**Silver Trauma Review Clinic**
**Patient Name:** **MRN :** **Date:**
**Assessor name and designation:**

| Falls Screen | Ax of Orthostatic Hypertension |
|---|---|
| **1.** In the past year have you had any other fall, including a slip or trip in which you lost your balance and lost your balance and landed on the floor or ground or lower level? **Y** ☐ **N** ☐<br>a. How many times did you fall in the past year? ________<br>b. How did you fall? (please describe e.g. activity, place, time):<br>C. Could you get up from the floor following your fall? **Y** ☐ **N** ☐<br>**2.** Are you afraid of falling? **Y** ☐ **N** ☐ Sometimes ☐ Don't know ☐<br>**3.** Did your fall require medical intervention? (specify):<br>**4.** Have you any difficulty with your walking or balance? (specify):<br>**5.** Do you get Dizzy or light headed? **Y** ☐ **N** ☐ Is this on standing up? **Y** ☐ **N** ☐<br><br>**ECG Done Y** ☐ **N** ☐ **Comments( specify):** | • **Lie down for 5 minutes. Take BP 1=**<br>• **Stand up. Take BP 2 in 1st minute=**<br>• **After 3 minutes take BP 3=**<br><br>**Symptoms (specify):**<br><br>POSITIVE RESULT **Y** ☐ **N** ☐<br>• **Drop in systolic BP of 20mmHG or more.**<br>• **A drop in diastolic BP of 10mmHg or more with symptoms.**<br>• **A drop below systolic 90mmHg on standing.** |

### PHYSIOTHERAPY MUSCULOSKELETAL ASSESSMENT

**Special Tests :**

**Walking: TUG (>20 seconds more detailed assessment) :**

**Balance: time held on one foot (<5seconds more detailed assessment):**

**Muscle strength:5xSTS (>15 seconds more detailed ax):**

**Subjective fear of falling: Y** ☐ **N** ☐

**Arom:**

**Power:**

**Balance:**

**Gait Assessment:**

**International Exercise Recommendations for older Adults:**

**Do you get 30 mins of aerobic exercise on any day during the week?**

**Do you do resistance training on any day during the week?**

**Do you do balance training on any day during the week?**

**Assessment Summary/ Treatment Plan**

| Silver Trauma Review Clinic | | |
|---|---|---|
| **Patient Name:** | **MRN :** | **Date:** |
| **Assessor name and designation:** | | |

| Action Plan | | |
|---|---|---|
| **Problems Identified** | **Actions** | **Completed** |
| **1.** | | |
| **2.** | | |
| **3.** | | |
| **4.** | | |

| Patient Follow up Plan of Care | | |
|---|---|---|
| **Interventions** | **Follow up** | **Referrals made** |
| | | GP ☐ |
| **Pain management** ☐ | | Geriatrician ☐ |
| **Immobilisation device** ☐ | Follow up in STR clinic Date……………… | Fracture Clinic ☐ |
| **Bone health** ☐ | | Integrated Care Team Older Persons (ICT) ☐ |
| **Medication alteration** ☐ | | Iv Zol clinic ☐ |
| **Falls prevention** | Discharged to GP ☐ | Others ☐ |
| | Discharged to Geriatric OPD ☐ | |
| Health Promotion ☐ | Discharged to IV Zol Clinic ☐ | |
| | Appointment made ☐ | |
| | Voice message left to Sec ☐ | |

| **Assessor name and designation:** «USERIMC» | **Date:** | **Time:** |
|---|---|---|

**Secondary Survey** **By Dr:**_______________________________________**@** **………… : ……….hrs**

**Silver Trauma Review Clinic**

**Patient Name:** **MRN :** **Date:**

**Assessor name and designation:**

| **Review #1** |
| --- |
| **Imaging/interventions:** |
| **Physiotherapy**: |
| **Advanced Nurse Practitioner**: |
| **Geriatrician** |
| **Trauma** |
| **Follow up plan:** |
